# Distinct clinicopathological differences between early gastric cardiac and non-cardiac carcinomas: A single-center retrospective study of 329 radical resection cases

**DOI:** 10.1101/2020.04.04.20052852

**Authors:** Yao Hui Wang, Xiu Qing Li, Li Li Gao, Chen Xi Wang, Yi Fen Zhang, Qin Huang

## Abstract

**Background:** Early gastric carcinoma is heterogeneous and can be divided into early gastric cardiac carcinoma (EGCC) and early gastric non-cardiac carcinoma (EGNCC) groups. At present, differences in clinicopathology remains obscure between EGCC and EGNCC fundus-corpus and antrum-angularis-pylorus subgroups, especially between EGCC with and without esophageal invasion.

**Methods:** In this study, we studied 329 consecutive early gastric carcinoma radical gastrectomies with 70 EGCCs and 259 EGNCCs.

**Results:** Compared to the EGNCC antrum-angularis-pylorus (n=181), but not fundus-corpus (n=78), sub-group, EGCC showed significantly older age, lower prevalence of the grossly depressed pattern, better tumor differentiation, higher percentage of tubular/papillary adenocarcinoma, but lower frequency of mixed poorly cohesive carcinoma with tubular/papillary adenocarcinoma, and absence of LNM in tumors with invasion up to superficial submucosa (SM1). In contrast, pure poorly cohesive carcinoma was less frequently seen in EGCCs than in EGNCCs, but mixed poorly cohesive carcinoma with tubular/papillary adenocarcinomas was significantly more common in the EGNCC antrum-angularis-pylorus sub-group than in any other group. No significant differences were found between EGCC and EGNCC sub-groups in gender, tumor size, *H. pylori* infection rate, and lymphovascular/perineural invasion. EGCC with oesophageal invasion (n=22), compared to EGCC without (n=48), showed no significant differences in the *H. pylori* infection rate and oesophageal columnar, intestinal, or pancreatic metaplasia, except for a higher percentage of the former in size > 2 cm and tubular differentiation.

**Conclusions:** There exist distinct clinicopathologic features between EGCC and EGNCC sub-groups; EGCC was indeed of gastric origin. Further investigations with larger samples are needed to validate these findings.

## Background

Gastric cancer is heterogenous in epidemiology, pathology, and pathogenesis mechanisms, and may be divided into cardiac and non-cardiac categories^1,2^, both of which show dismal prognosis. At present, the best strategy to improve gastric cancer patient outcomes is early detection with prompt resection of early gastric carcinoma (EGC). Once EGC is diagnosed, endoscopic therapy, such as endoscopic mucosal resection and endoscopic submucosal dissection, has gradually replaced conventional surgical resections in selected cases because of fewer complications, better post-resection recovery, and lower hospital cost. Section criteria for endoscopic therapy is mainly based on the risk of lymph node metastasis (LNM) in patients with EGC^3^. However, it remains elusive about the differences in the risk of LNM between early gastric cardiac carcinoma (EGCC) and early gastric non-cardiac carcinoma (EGNCC). Previous studies have shown that advanced gastric cardiac cancer may have clinicopathological features and pathobiological behaviors distinctly different from gastric non-cardiac cancer because of higher pT and pN stages and poorer prognosis^4,5^. We hypothesized that these differences may be related, at least in part, to various mucosal epithelial cell types housed in 3 different gastric regions: 1. cardia with predominant cardiac and cardiofundic mucosa composed of mainly mucinous and some mixed oxyntic glands; 2. fundus and corpus with fundic-type mucosa equipped with oxyntic glands; and 3. antrum-angularis-pylorus with antral-type mucosa consisting of mucinous glands. As such, EGC from these 3 different regions of the stomach may have discrete clinicopathologic features and various risks of LNM, which however, have not been studied, to the best of our knowledge. Thus, the aims of the present study were to investigate clinicopathology and risk of LNM in EGC arising in the gastric cardia, fundus-corpus, and antrum-angularis-pylorus regions in patients treated at our center in the Jiangsu Providence, which is one of gastric cancer endemic regions in China.

## Methods

### Patient selection

We searched the electronic pathology database stored in the Jiangsu Province Hospital of Chinese Medicine in Nanjing, China, over the 7-year period from January 2011 to December 2017 for gastric cancer radical resection and collected 2184 consecutive cases with pathologically confirmed gastric cancer. According to the 2019 World Health Organization (WHO) diagnostic criteria^6^, 443 EGC cases were identified. Two experienced pathologists reviewed histological slides of all EGC cases, 114 of which were excluded because of stump carcinoma (n=11), synchronous carcinoma (n=2), high-grade intraepithelial neoplasia (n=27), lymphoma (n=2), and neoadjuvant chemotherapy (n=72). As a result, 329 EGC cases were eligible for this study. The clinicopathological characteristics of all cases, based on pathologic and endoscopic reports and operative notes, were tabulated and analyzed, which included gender, age, tumor location, size, and gross patterns. Each case was pathologically staged, according to the 8^th^ edition of the American Joint Committee on Cancer staging manual^7^. In this study, all patient’s private information was deleted to protect the patient’s privacy. The study protocol was approved by the hospital Medical Ethics Committee (Number: 2019NL-098-02).

### Pathologic study

The 5th edition WHO diagnostic criteria on gastric carcinoma were followed and EGC was defined as invasive carcinoma confined to the mucosa or submucosa^6^. EGCC referred to the tumor with its epicenter located within 3 cm below the gastroesophageal junction (GEJ)^8^. For EGNCC, the tumor epicenter was situated in the distal stomach, more than 3 cm below the GEJ either in the fundus-corpus region of the stomach or more distally in the gastric antrum-angularis-pylorus region. Tumor gross features were divided into 5 subgroups: type 0-I (polypoid/protruding), type 0-IIa (superficial elevated), type 0-IIb (flat), type 0-IIc (superficial depressed), and type 0-III (excavated)^6^. These 5 sub-subgroups were further simplified into 3 sub-groups: Types 0-I and 0-IIa were sub-grouped as the elevated type; Type 0-IIb as the flat type, and Types 0-IIc and 0-III as the depressed type, for a simplified statistical analysis. The depth of tumor invasion was divided into 4 sub-groups as follows: M2 (tumor infiltration confined to the lamina propria without the involvement of the muscularis mucosae), M3 (tumor invasion into muscularis mucosae), SM1 (tumor involvement of submucosal superficial layer with the infiltration depth < 500 μm from the muscularis mucosae), and SM2 (tumor involving the submucosal deep layer with the infiltration depth ≥ 500 μm from the muscularis mucosae). Lymphovascular invasion and perineural invasion were also recorded. In equivocal cases, routine elastic fiber staining and immunostaining for CD31 and D2-40, with valid controls, were carried out to validate the finding of lymphovascular invasion recognized on conventional hematoxylin-eosin stains. Guided by the WHO diagnostic criteria^6^, tumor differentiation (well, moderate, or poor) and histopathological type (tubular adenocarcinoma, papillary adenocarcinoma, micropapillary adenocarcinoma, poorly cohesive carcinoma [PCC] including signet-ring cell carcinoma, mucinous carcinoma, mixed PCC and tubular/papillary adenocarcinoma, mixed mucinous and tubular/papillary adenocarcinoma) of EGC were tabulated and analyzed. *Helicobacter pylori* (HP) infection was confirmed by microscopic identification of the bacterium on routine hematoxylin-eosin and basic fuchsin stains with appropriate controls.

In most Chinese patients, endoscopic mucosal GEJ and squamous-columnar epithelial junction lines overlap at the same level ^9^. Microscopically, the histologic criteria for the GEJ line were defined as the distal end of oesophageal squamous epithelium, multilayered epithelium or oesophageal submucosal glands or ducts^10^. Once the GEJ line was identified, the tumor epicenter location and the extend of tumor invasion were determined; the distance of oesophageal invasion was measured microscopically with the assistance of an ocular scale bar. In addition, distal oesophageal columnar metaplasia, intestinal metaplasia, pancreatic metaplasia, and dysplasia were also studied and analyzed.

### Statistical analysis

Differences between groups with continuous or categorical variables were statistically compared with appropriate statistical methods, such as Student t, χ2, Fisher’s exact, or Kruskal-Wallis H test. *P* values < 0.05 were considered statistically significant. All statistical analyses were performed with the All analyses were performed using SPSS software, version 13.0 (IBM, Armonk, NY).

## Results

As shown in Table 1, among 329 consecutive eligible EGC cases, 70 were classified as EGCC (21.9%, 70/329) and 259 (78.1%, 259/329) were as EGNCC. In the EGNCC group, 78 (24.5%, 78/329) were in the fundus-corpus region and 181 (56.7%, 181/329) were in the Antrum-angularis-pylorus region. Overall, the number of male patients was predominant in all groups but the difference in gender was not statistically significant. In contrast, the average age of patients was 60.2 years (range: 18-83) for the cohort and significantly much more advanced in EGCC than in other sub-groups (*P* < 0.01).

**Table 1.**
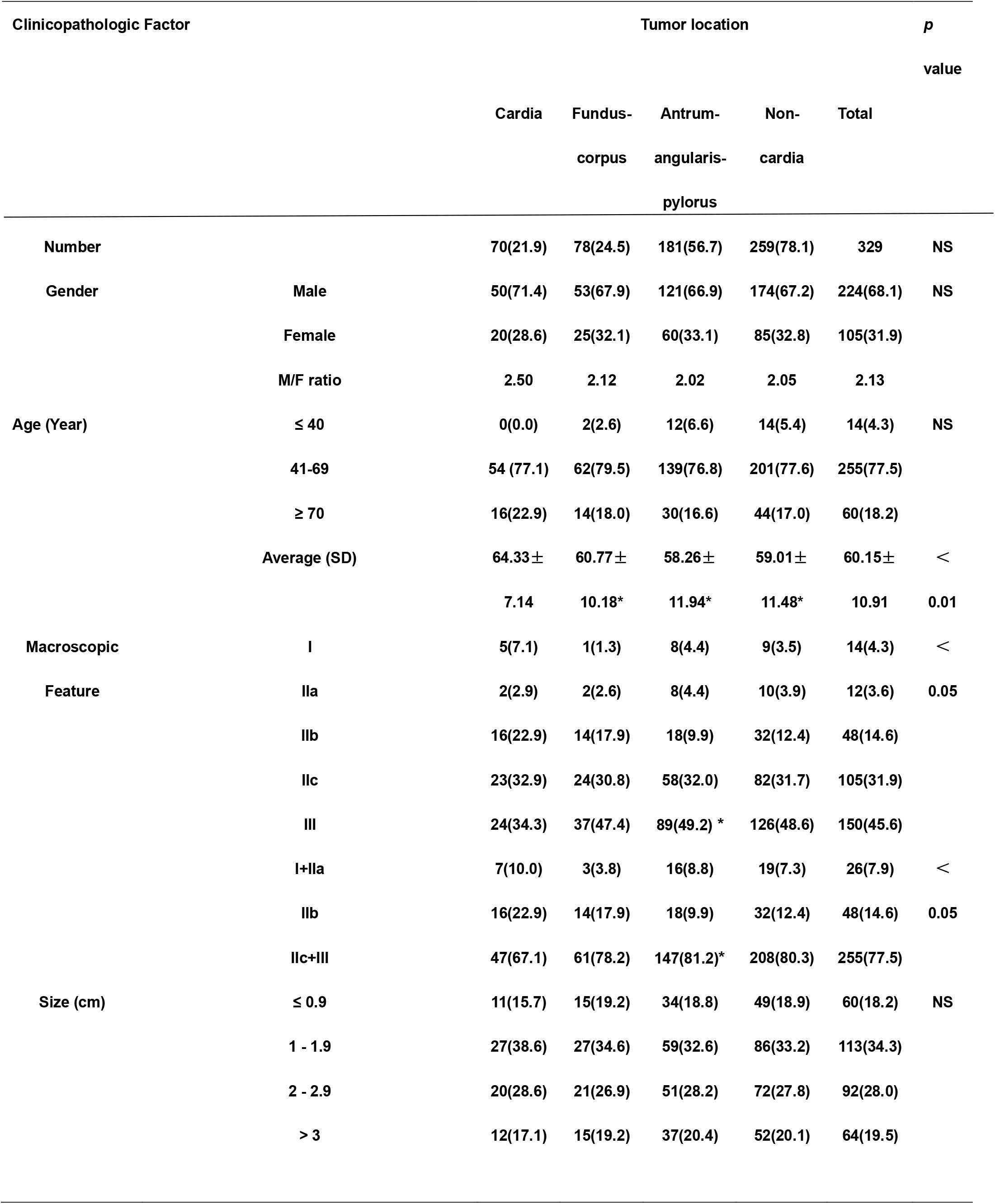
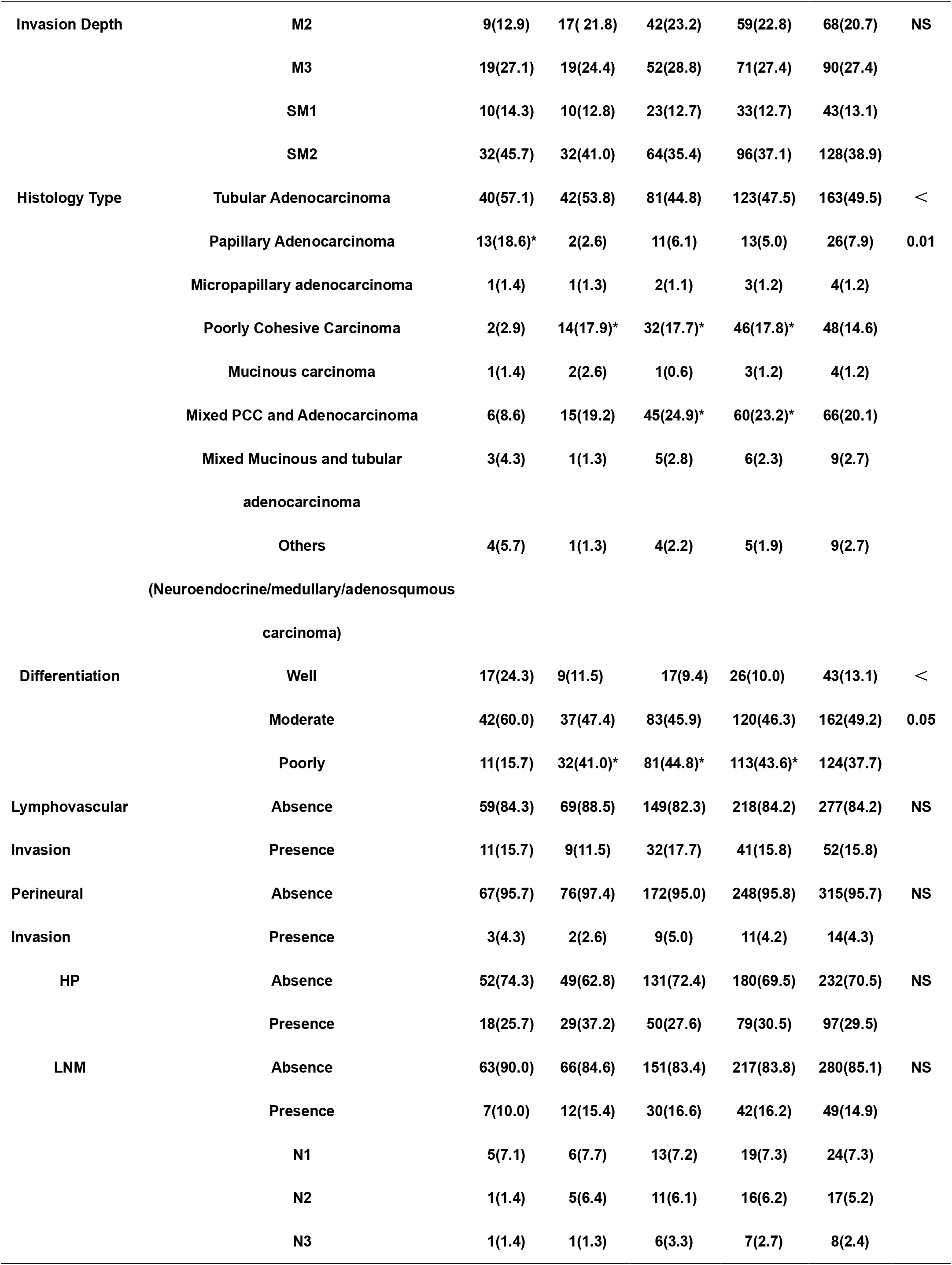
**Comparison of Clinicopathologic Features of EGC among Different Locations**

### Macro- and microscopic differences in pathology between early gastric cardiac and non-cardiac carcinomas

As shown in Table 1, while overall differences in macroscopic growth patterns of EGC were significant between EGCC and EGNCC groups (*P* < 0.05), there were no significant variations in the elevated or flat sub-group of EGC between the two groups, despite the fact that the frequency of the elevated pattern was more commonly seen in the former (10.0%) than in the latter (7.3%). However, EGC tumors with the depressed pattern (Types 0-IIc and 0-III) were significantly less commonly seen in the EGCC group (67.1%) than in the antrum-angularis-pylorus group (81.2%) (*P* < 0.05), but not in the fundus-corpus sub-group of ENGCC tumors. Statistically, differences in overall tumor size and invasion depth were not significant between EGCC and ENGCC sub-groups.

In this cohort, differences in the EGC histologic type were significantly different between EGCC group and EGNCC sub-groups (*P* < 0.01). As displayed in Table 1, tubular and papillary **(Fig.1A)** adenocarcinomas were more frequently seen in the former than in the latter, especially for papillary adenocarcinoma that was significantly more prevalent in EGCC (18.6%) than in EGNCC (5.0%) (*P* < 0.05). In contrast, pure PCC **(Fig.1B)** was significantly less frequently observed in EGCC than in EGNCC fundus-corpus and antrum-angularis-pylorus sub-groups (*P* < 0.05), while mixed PCC with tubular/papillary adenocarcinoma was more commonly seen in the antrum-angularis-pylorus (*P* < 0.05), but not the fundus-corpus, sub-group of EGNCC. There were no significant differences in uncommon variants of EGC, such as micropapillary adenocarcinoma **(Fig.1C)**, pure and mixed mucinous adenocarcinomas, neuroendocrine and adenosquamous carcinomas, and carcinoma with lymphoid stroma, between EGCC and EGNCC sub-groups. In the cohort, tumor differentiation was significantly different (*P* < 0.05) between the EGCC and EGNCC groups. Although well differentiation in EGC was more commonly noted in EGCC (24.3%) than in EGNCC (10.0%) groups, the difference did not reach a statistically significant level. However, poor differentiation in EGC was significantly less commonly identified in EGCC (15.7%) than in EGNCC (43.6%) groups (*P* < 0.05). There were no significant differences between EGCC and EGNCC groups in lymphovascular invasion, perineural invasion, HP infection rate, and LNM.

**Figure 1:**
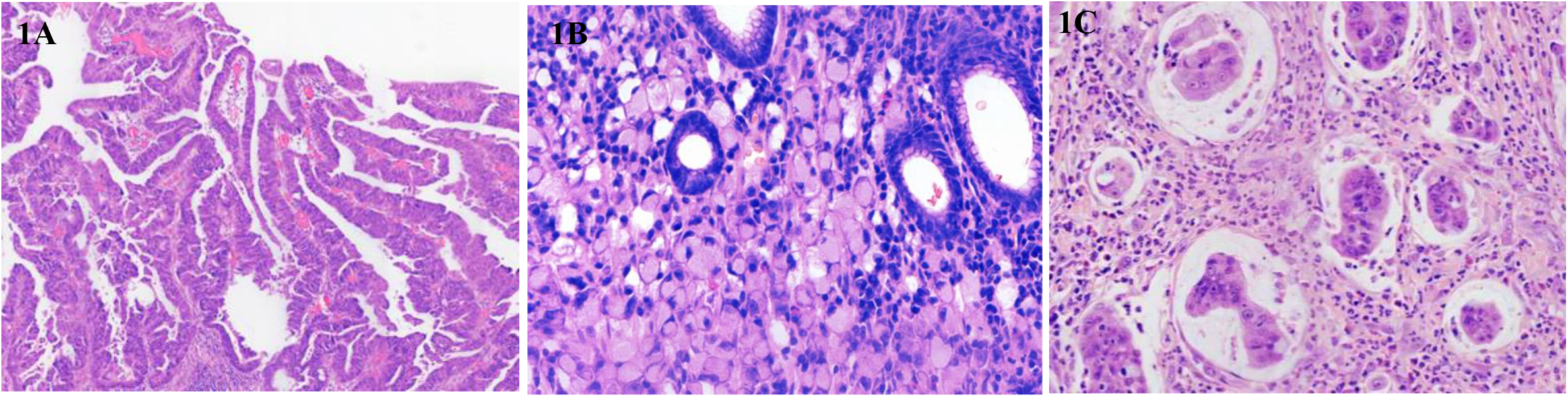
Representative images of papillary (A), signet ring (B) and micropapillary (C) early gastric cardiac carcinomas.

### Depth of invasion and lymph node metastasis

The average number of regional lymph nodes retrieved and examined per case was 21.5 (range: 2 to 68). As illustrated in Table 2, the prevalence of LNM in the cohort was 5.7% (9/158) for intramucosal carcinoma and significantly increased to 23.4% (40/171) for submucosal carcinoma (*P* < 0.01). Surprisingly, no LNM was detected in 38 EGCC cases with the invasion depth up to superficial submucosa, ie, M2+M3+SM1; thus, the prevalence of LNM was significantly lower in EGCC than in EGNCC (9.2%, 15/163) groups (*p* < 0.05). Specifically, the percentage of LNM in EGNCC was 6.8% (4/59), 7.0% (5/71), and 18.2% (6/33) for invasion depth at M2, M3, and SM1, respectively. There was no significant difference in risk of LNM of EGC with deep submucosal invasion (SM2) between EGCC and EGNCC sub-groups.

**Table 2.**
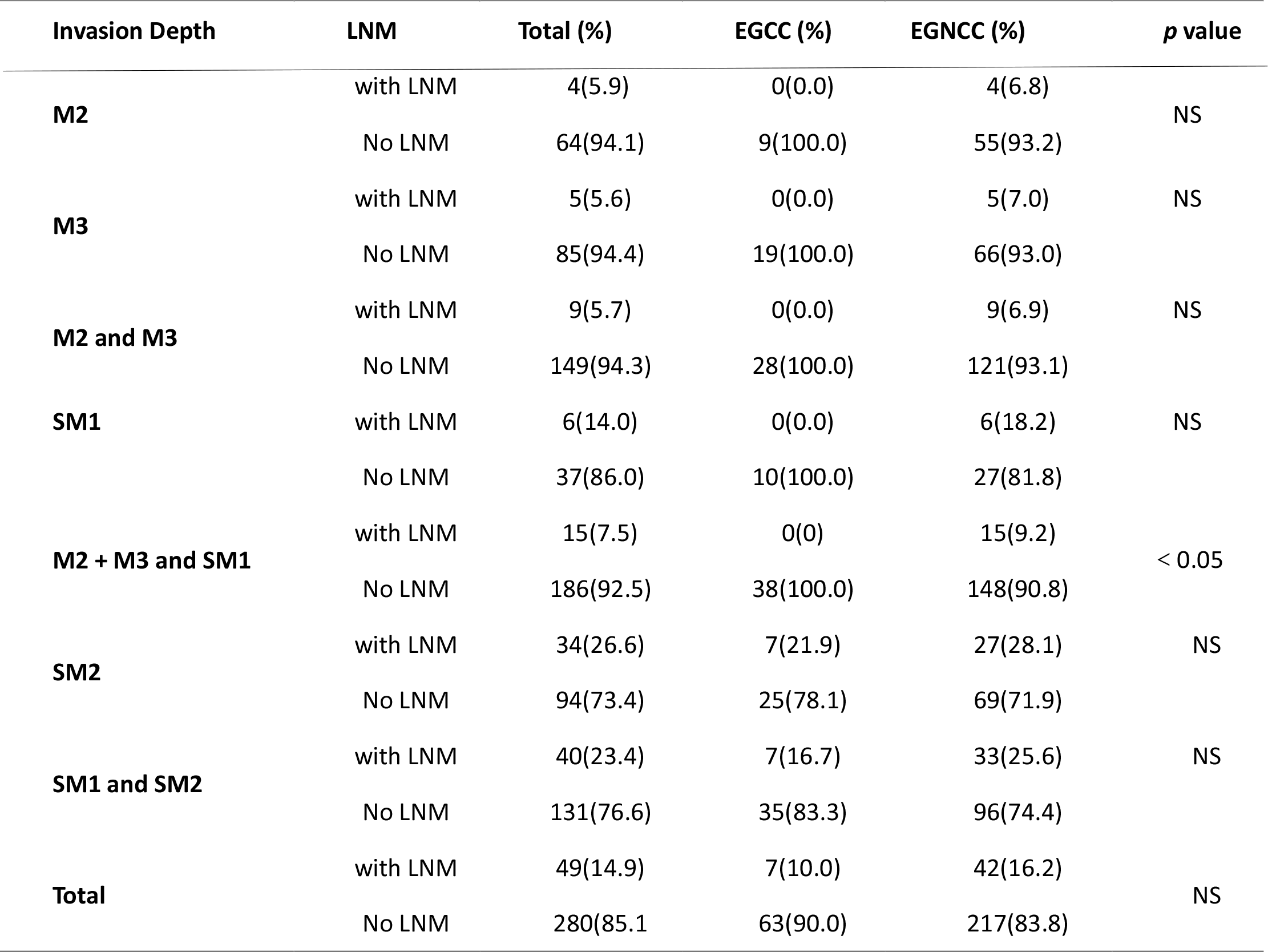
**Comparison of Relationship of Invasion Depth and Lymph Nodal Metastasis of Different Sites**

### Early gastric cardiac carcinoma with oesophageal invasion

In this study, 22 (31.4%, 22/70) EGCC tumors showed their epicenters in the gastric cardia with a small proportion of the tumor crossing the GEJ line into the distal oesophagus **(Fig.2A)**, extending to an average distance of 4.16 mm (range: 1-10), primarily underneath benign oesophageal squamous epithelium. No Barrett’s oesophagus was recognized in all 22 EGCC cases, in addition, high-grade intraepithelial neoplasia was observed in the pericancerous mucosa **(Fig.2B)**, suggesting that EGCC originated in the stomach. As demonstrated in Table 3, epithelial columnar metaplasia in the distal oesophagus was seen in 50 (71.4%, 50/70) cases, in which 34 were columnar mucinous metaplasia (68.0%, 34/50), 13 were pancreatic metaplasia (26.0%, 13/50), and only 3 cases showed intestinal metaplasia (6%, 3/50). No epithelial dysplasia was identified in metaplastic epithelium. Compared to EGCC cases without oesophageal involvement (68.6%, 48/70), EGCC cases with oesophageal invasion showed no significant differences in gender, age, tumor origin, epithelial metaplasia, tumor macroscopic pattern, overall size, invasion depth, differentiation, lymphovascular or peri-neural invasion, and LNM. Interestingly enough, the prevalence of cases with tumors larger than 2 cm in size was significantly more frequently present in EGCCs with oesophageal invasion (68.2%, 15/22) than those without (35.4%, 17/48) (*P* < 0.05) and the overall difference in the microscopic tumor type was also statistically significant between the two sub-groups (*P* < 0.01), in that EGCC tumors with oesophageal invasion showed a significantly lower proportion of tubular adenocarcinoma than those without oesophageal invasion.

**Figure 2:**
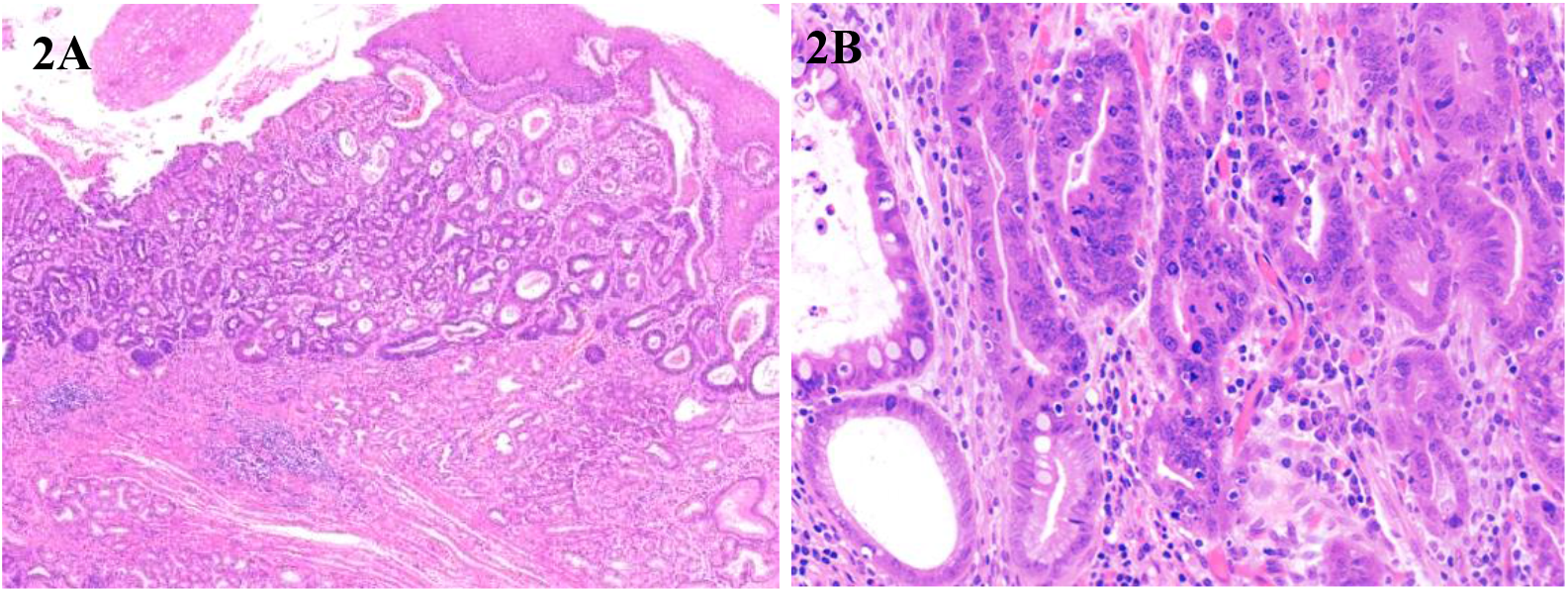
Early gastric cardiac carcinoma with oesophageal invasion. A: A small component of early gastric cardiac carcinoma invades into the distal oesophagus between superficial oesophageal glands and squamous epithelium. B: The high-grade intraepithelial neoplasia in the pericancerous mucosa was showed.

**Table 3.**
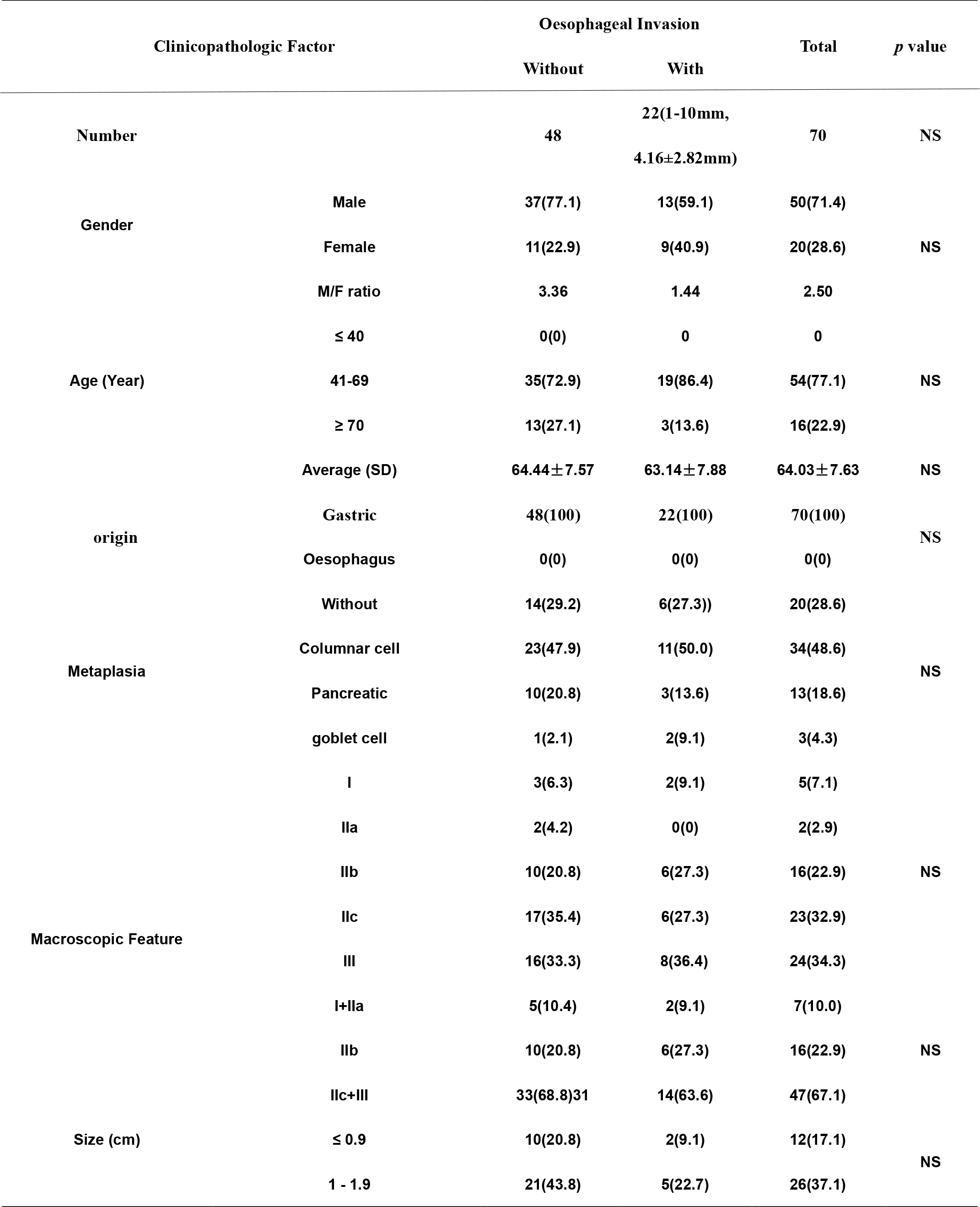
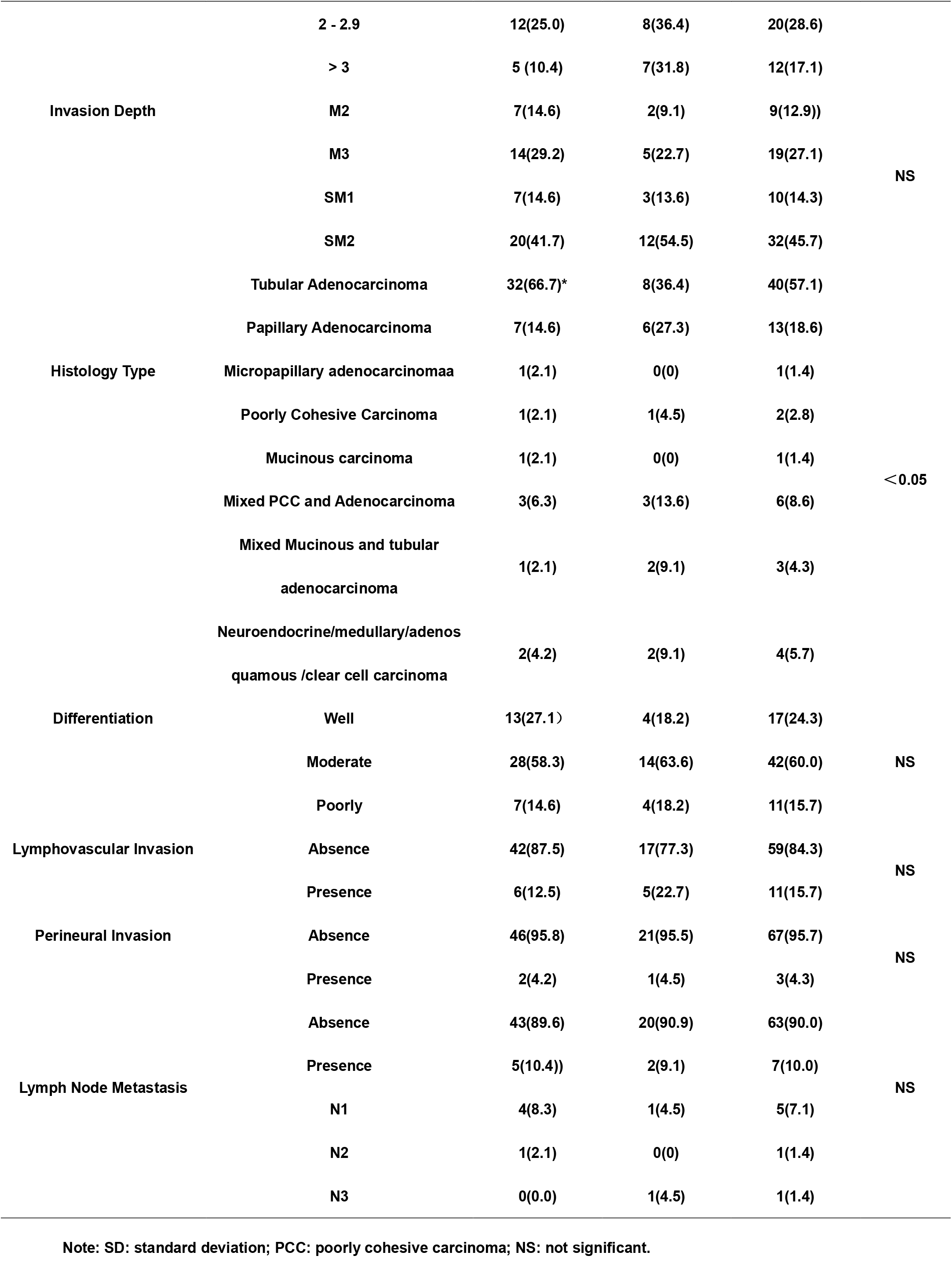
**Comparison of Clinicopathologic Factors of Early Gastric Cardiac with or without oesophageal Invasion**

## Discussion

In this single-center clinicopathology study, we showed several important features in EGCC, compared to those in EGNCC subgroups, including more advanced patient age, better tumor differentiation, higher prevalence of tubular, especially papillary adenocarcinomas, but lower percentages of poor tumor differentiation and PCC. No significant differences were observed in gender, HP infection rate, tumor size, lymphovascular/perineural invasion between the two groups, but LNM in EGCC with invasion up to SM1 was absent. Importantly, we showed no significant differences in the HP infection rate, the prevalence of intestinal metaplasia, and the risk of LNM between EGCC with and without oesophageal invasion sub-groups. Moreover, intestinal metaplasia in the distal oesophageal columnar metaplastic epithelium remained rare (6%) in EGCC cases with oesophageal invasion. These findings, if confirmed with larger samples in the future, may have lasting impact on the current clinical management strategy for patients with EGC, especially EGCC.

EGC heterogeneity is well-known in risk factors, histopathology, molecular pathobiology, and prognosis. The percentage of EGCC in our cohort was much higher than that previously reported in Japan^11,12^, Korea^13^, and The United States^14^, but similar to those reported in China^15,16^, illustrating marked geographic variations in this cancer. By histopathology, the vast majority of EGCC tumors in our study were tubular and papillary adenocarcinomas, but PCC was uncommon, which is in sharp contrast to EGNCC, as we previously reported^15,16^. These morphologic differences between EGCC and EGNCC are supported by various genomic types of gastric carcinoma in that the chromosomal instable-type gastric carcinoma, mainly manifested as tubular/papillary adenocarcinoma, is much more common, but the genomic stable variant, such as PCC, is much less frequent in the gastric cardia, compared to distal gastric regions^17^. We did not observe significant histopathologic differences between EGNCC fundus-corpus and antrum-angularis-pylorus subgroups. This may be related, in part, to small sample sizes of cases in the current study and await further investigation with larger samples in the future.

While HP infection has been shown to be the most important risk factor for EGC and inversely related to Barrett’s adenocarcinoma^18^, we did not see significant differences in the HP infection prevalence between EGCC and EGNCC, as we reported previously^15,16,19^, suggesting an important role of the HP infection in EGCC pathogenesis. Apparently, the pathogenesis mechanisms of EGCC differ from those of Barrett’s adenocarcinoma but more like those of gastric carcinoma. On the other hand, advanced age was again revealed to be significantly more commonly observed in EGCC than in EGNCC, as reported previously^15,16,19^. Although detailed mechanisms on aging-related tumorigenesis for EGCC remain unknown, these results suggest different pathogenesis pathways between EGCC and EGNCC.

Compared to EGNCC, EGCC tumors invaded deeper with a higher percentage of cases with submucosal invasion, as we reported recently in a multicenter study^20^. Submucosal EGC has been shown to have aggressive behaviors with high risk for LNM^16,21^. In our study, this is true for EGNCC, but not for some EGCC cases, in which no LNM was observed in EGCC cases with invasion to superficial submucosal (SM1). This finding may be related to small samples with SM1 EGCC, but the result may at least suggest a lower risk of LNM in SM1 EGCC and lent support to the role of endoscopic therapy for intramucosal and some qualified superficial submucosal EGCC tumors^20^. EGCC pathologic staging stays controversial. If tumor epicenter is located within 2 cm of the GEJ line with invasion into distal oesophagus, EGCC is required to be classified and staged as oesophageal carcinoma by the 8^th^ edition of the American Joint Committee on Cancer staging manual^7^. This decision assumed that gastric cardiac carcinoma with oesophageal invasion might be a part of Barrett’s adenocarcinoma^22^. This appears to be debatable. In our cohort, HP infection in EGCC was common but intestinal metaplasia in the oesophageal columnar metaplastic epithelium was rare, unlike in Barrett’s adenocarcinoma. In addition, ogastroesophageal reflux disease, hiatal hernia, and Barrett’s oesophagus and adenocarcinoma continue to be scarce in our study patient population. Furthermore, we did not discover significant differences in gender, age, oesophageal columnar/intestinal metaplasia, and LNM between EGCC with and without oesophageal invasions. These lines of evidence argue against the classification and staging of EGCC with focal oesophageal invasion as oesophageal carcinoma.

Major limitations of this study include the retrospective study design with no standardized lymphadenectomy so that the number of lymph nodes retrieved varied. However, the overall quality of nodal retrieval remained sound because of a high average number of lymph nodes (over 21) retrieved per case in this cohort. The other undesirable issue in this study was the small number of EGC cases in 3 sub-regions: cardia, fundus-corpus, and antrum-angularis-pylorus so that differences in several clinicopathologic variables could not be effectively analyzed statistically. To overcome this short-coming, our next research plan was to expand the current investigation into a multi-center study, which is on-going.

In summary, EGCC, compared to EGNCC, showed more advanced patient age, better tumor differentiation, higher percentage of tubular and papillary adenocarcinomas, but lower frequency of PCC, and lower risk of LNM. EGCC with epicenter in the gastric cardia may invade the oesophagus, but exhibited rare intestinal metaplasia and high prevalence of HP infection in our cohort, which support the classification of EGCC as gastric, not oesophageal, carcinoma. Further investigations with larger samples in multi-center studies are urgently needed to validate our findings for a better understanding of EGCC and more appropriate patient management strategy.

## Data Availability

The raw data generated and analyzed in the current study are not publicly available due to appropriate protection of patient personal information but are available from the corresponding author on reasonable request.

## Abbreviations

EGC: Early gastric carcinoma
EGCC: Early gastric cardiac carcinoma
EGNCC: Early gastric non-cardiac carcinoma
LNM: Lymph node metastasis
WHO: World health organization
GEJ: Gastroesophageal junction
PCC: Poorly cohesive carcinoma
HP: Helicobacter pylori

## Declarations

### Ethics approval and consent to participate

This study is a retrospective analysis, the study protocol was approved by the hospital Medical Ethics Committee (Number: 2019NL-098-02).

### Consent for publication

Not applicable

### Competing interests

The authors declare that they have no competing interests.

### Funding

This work was supported by clinical study initiated by researchers at Jiangsu Province Hospital of Chinese Medicine (YJZ201901). The funding bodies played no role in the design of the study, the collection, analysis and interpretation of data or in the writing of the manuscript.

### Authors’ contributions

WYH analyzed the data and wrote the initial draft of the manuscript. LXQ, WCX collected pathological data. GLL was responsible for pathological technique operation. HQ, ZYF critically appraised and revised the overall content of the study and manuscript. All authors read and approved the final manuscript.

#### Acknowledgements

Not applicable

## Conflict of interest

The authors have no conflicts to disclose.

## Notes

### Competing Interest Statement

The authors have declared no competing interest.

